# Summarizing data from continuous glucose monitors using the cgmstats package

**DOI:** 10.64898/2026.03.30.26349753

**Authors:** Natalie Daya Malek, Dan Wang, Sui Zhang, Michael Fang, Amelia Wallace, Scott Zeger, Elizabeth Selvin

## Abstract

In this article, we present the **cgmstats** package for the analysis of continuous glucose monitoring (CGM) data. The use of wearable CGMs is growing rapidly. The latest generation of CGM systems do not require fingerstick calibration, are minimally invasive, and are frequently used in research studies. CGM sensors are typically worn for up to 2 weeks and record interstitial glucose measurements every minute to every 15 minutes, depending on the sensor used. CGM systems generate hundreds of measurements per day and thousands of measurements in one person over a single wear. There is a need for tools that allow researchers to efficiently organize and summarize the wealth of data on glucose patterns produced by CGM systems. The **cgmstats** package generates CGM summary measures for data from a variety of CGM systems and allows the user to flexibly define ranges and generate data visualizations. In this article, we provide an overview of the **cgmstats** package and examples of its use. The **cgmstats** package supports rigorous and reproducible analyses of CGM data.

## 1. Introduction

CGMs were originally designed for the clinical management of type 1 diabetes. However, their use has expanded rapidly, and these devices are now used by patients with type 2 diabetes and even individuals without diabetes. There have been numerous randomized clinical trials demonstrating the benefits of CGM use for managing glucose control (Beck et al. 2017; Martens et al. 2021), and CGMs have been incorporated into large, population-based studies to examine glucose patterns in the general community and their associations with clinical outcomes (Daya et al. 2024; Shah et al. 2019; Selvin et al. 2021; Spartano et al. 2025). CGM systems generate a large amount of data (i.e., ~1,300 to ~21,000 readings over 15 days depending on the sensor) in different formats. Flexible and reproducible tools are needed for researchers to analyze and interpret these data. While CGM manufacturers produce CGM summary reports for patients and their providers, these reports are designed for clinical use and typically summarize up to ~2 weeks of data, on a single person, with a limited number of CGM summary statistics.

There is currently no standardized approach to the processing or analysis of CGM data. This lack of standardization in the field is complicated by the diversity of data output formatting across CGM systems, making it difficult to directly compare results including CGM summary statistics across studies. Thus, there is a need for flexible statistical packages that can process a wide variety of data and conduct analyses using different approaches. Examples of key aspects of flexibility needed include: being able to vary cut points for time in range and time above and below range; the ability to alter the definition of hypoglycemic or hyperglycemic episodes based on level and duration; requiring a certain percentage of valid readings (e.g., 70%) for the summary metrics to be considered reliable; and being able to drop the first or last 24 hours of data. Typically, CGM sensor data are considered less reliable during the first 24 hours of wear. During this time, a CGM sensor may have unexpectedly high or low readings as it is initializing and calibrating. Over time, sensor degradation or changes in adhesion to this skin may lead to inaccurate readings or signal interruptions towards the end of the wear period. There may be also gaps in the data due to issues with the sensor or transmitter (i.e. sensor displacement, poor adhesion, software glitches). There is no standard approach to how to handle these issues.

In prior studies, including major randomized clinical trials, it is unclear if some CGM data points were excluded or if all available sensor readings were used (Aleppo et al. 2017). For example, some studies drop the first and last day of CGM data but others do not (Beck et al. 2017; Wan et al. 2018). The handling of missing data is also variable. While some studies impute small gaps (i.e., less than 30 minutes) using interpolation methods, others do not (Kuang et al. 2024). Large gaps may lead to exclusion of participants if the missing data exceeds a predefined threshold to deem the summary statistic reliable (i.e., the guidelines suggest a minimum of 70% of data is captured from the total CGM wear time).

The lack of standardization in data processing methods across studies including clinical trials involving CGM data suggests a need for clearer reporting and methodological consistency to enhance the reliability of findings.

Automating the calculation of CGM metrics is efficient and ensures an accurate and reproducible approach. Several packages exist, primarily in R or as web-based or desktop applications, to process and summarize CGM data (Karakus, Snell-Bergeon, and Akturk 2024). To our knowledge, there are no Stata software packages that provide a flexible and automated approach to process and comprehensively analyze CGM data. Also, existing packages often lack the flexibility to accommodate data from the more than 10 major CGM systems used globally which store glucose readings at various intervals. They also lack the flexibility to allow users to easily drop a specified number of 24-hour periods from the start or end of the wear period or restrict the analysis to a specified period according to dates and times, all in one package. Most software packages do not automatically generate individual time-series data for a given person to examine how an individual’s CGM summary metrics vary by day.

We created the first Stata package to process, summarize, and visualize data from CGM systems. **cgmstats** processes data obtained from different CGM systems containing 3 columns of data (ID, date/timestamp, and CGM glucose reading) and outputs a Stata dataset with a variety of CGM summary metrics, including standard clinical summary measures used in the management of diabetes. These include percent time in range, time spent at high (hyperglycemia) or low levels of glucose (hypoglycemia), the count and duration of hypo- and hyperglycemic episodes, and measures of glycemic variability (Battelino et al. 2023; American Diabetes Association Professional Practice 2025).

The **cgmstats** package provides user flexibility to analyze CGM data from a variety of sensors and different generations of devices from the same manufacturers, to define thresholds and duration of hypo- and hyperglycemic episodes, and output CGM summary metrics into a new dataset by person or by day, or as a time-series dataset by person and day. The package accepts CGM data with gaps. It also provides the user capability to visualize CGM glucose tracings over the wear period with the option to fill in gaps in the data using linear interpolation.

As CGMs are increasingly used by patients and in research studies, there is a growing need for software to summarize and visualize CGM time series using a flexible approach and with automated output of standard CGM metrics consistent with clinical practice. The **cgmstats** package fulfills this need in Stata.

## 2. The cgmstats package

### 2.1 Description

The **cgmstats** package processes the CGM data according to the user’s specifications and creates a dataset with summary CGM metrics. By default, the package generates measures using definitions recommended in clinical guidelines (Battelino et al. 2023). Users also have the option to create CGM metrics using custom cut points. The package allows users to define the period of analysis and includes functions to exclude biologically implausible values (e.g. low values caused by pressure placed on the sensor during sleep; high or low oxygen tensions; improper sensor calibration). The summary CGM metrics are calculated overall and separately by daytime and nighttime. The package accepts glucose measurements in units of mg/dL (default) or mmol/L and outputs metrics in the same unit.

The **cgmstats** package produces and saves histograms of user-specified CGM summary metrics. It also plots CGM tracing per person if specified by the user.

### 2.2 Syntax

The input Stata dataset(s) must contain 3 columns with the following information: ID (unique identifier for each person), timestamp (date and time of glucose reading), and sensor glucose reading. The user specifies the variable name for each of the 3 columns.

Raw CGM data is typically received in the comma-separated values format. The user may prepare the respective .dta files using the command `-import excel-`.

All input Stata datasets should be saved in the user-specified directory or if not specified in the package syntax, the working directory (by default).

The package syntax is:

~~~
cgmstats, id(*varname*) glucose(*varname*) time(*varname*) ///
dtadir(*string*) [freq(*#*) unit(*string*) ///
by(*string*) keep(*string*) ///
hyper(*numlist*) hypo(*numlist*) hyper_exc_lngth(*#*) ///
hypo_exc_lngth(*#*) daystart(*string*) dayend(*string*) ///
firsthours(*#*) lasthours(*#*)///
timebefore(string) timeafter(*string*) ///
auc hist(*varlist, [freq bw(#)]*) ///
plot(numlist, [nostats fill]) ///
savecombdta(*string*) savecombdir(*string*) ///
savesumdta (*string*) savesumdir(*string*) ///
saveplotdir(*string*)]
~~~

where *id(varname)* is the unique identifier (numeric or string variable) of the patient or participant, *glucose(varname)* is the numeric CGM glucose values and *time(varname)* is the timestamp of CGM measurement. The timestamp must be in the format %tcCCYY/NN/DD_HH:MM (e.g. “2014/12/01 01:30”). dtadir(*string*) specifies the directory containing all input files in dta format.

If the timestamp is in the incorrect format, the user will receive an error that says, for example:

~~~
Timestamp is not in the correct format %tcCCYY/NN/DD_HH:MM.
Current format: %tcCCYY-NN-DD_HH:MM:SS
~~~

Additionally, if duplicate timestamps are found, the user will receive the following error:

~~~
Duplicate timestamps detected!
~~~

If the directory containing the input dta files is not specified, the user will receive the following error:

~~~
You must specify the directory with the input dta-files
~~~

### 2.3 Options

**freq(***#*) establishes CGM recording frequency (in minutes), default is 15 minutes

**unit(**mmol/L) specifies glucose units are in mmol/L, default is mg/dL

**by(***day***) or by(***id day*) produces CGM metric summary files (datasets and plots) by day of CGM wear (i.e., by(day)), or produces one CGM metric summary file by ID and day of wear (multiple rows per ID) (i.e., by(id day))

**keep(***string*) contains conditional statement(s) to subset data according to glucose values or ID (i.e., keep(if sensorglucose>50))

**hyper(***numlist*) defines glucose value cut point(s) for a hyperglycemic episode. An episode is defined as no values below glucose value cut point(s) for a certain period of time. Default values are 140 mg/dL (7.8 mmol/L), 180 mg/dL (10 mmol/L) and 250 mg/dL (13.9 mmol/L). If unit(mmol/L) is specified, then cut points should be in units of mmol/L. Decimals are accepted but decimal points will be converted to “_” in variable names.

**hypo(***numlist*) defines glucose value cut point(s) for a hypoglycemic episode. An episode is defined as no values above glucose value cut point(s) for a certain period of time. Default values are 54 mg/dL (3 mmol/L) and 70 mg/dL (3.9 mmol/L). If unit(mmol/L) is specified, then cut points should be in units of mmol/L. Decimals are accepted but decimal points will be converted to “_” in variable names.

**hyper_exc_lngth(***#*) defines length (in minutes) of a hyperglycemic episode, default is 30 minutes. During this period, there are no values below X (as defined by option hyper(numlist)).

**hypo_exc_lngth(***#*) defines length (in minutes) of a hypoglycemic episode, default is 30 minutes. During this period, there are no values above X (as defined by option hypo(numlist)).

**daystart(***string*) defines the time indicating start of day (HH:MM, 24-hour scale), default is 06:00

**dayend(***string*) defines the time indicating end of day (HH:MM, 24-hour scale), default is 22:00

**firsthours(***#*) defines the number of hours to drop from the start of the CGM wear period. Accepts decimals (i.e., 0.5 for 30 minutes).

**lasthours(***#*) defines the number of hours to drop from the end of the CGM wear period. Accepts decimals (i.e., 0.5 for 30 minutes).

**timebefore(***timestamp*) subsets data to time before specified timestamp, i.e. timebefore(“2021/09/01 12:00”)

**timeafter(***timestamp*) Subsets data to time after specified timestamp, i.e. timeafter(“2023/06/01 12:00”)

**auc** calculates the AUC over glucose value cut point(s) defined by option hyper(numlist)

**hist(***string, [freq bw(#)]*) plots histograms of user-specified CGM summary statistics overlaid with kernel density plots [i.e., hist(mean_sensor cv_sensor)] by default. If option “*freq*” is used, the frequency histograms are plotted instead, without kernel density plots [i.e., hist(mean_sensor, freq)]. “*bw*(*#*)” executes the option to alter the bar widths of the histogram.

**plot(***numlist, [nostats fill]*) plots CGM glucose tracings over the entire CGM wear period with key CGM summary metrics (can be turned off using option “nostats”, i.e., plot(4 10, nostats). Users define the two cut points (low and high) for time in range and these cut points must be contained in options hypo(*numlist*) and hyper(*numlist*). If a decimal was specified in options hypo(*numlist*) or hyper(*numlist*), it must be rounded to nearest whole number for the plot option. “*fill*” executes option to fill in gaps in CGM glucose tracings using linear interpolation.

**savecombdta**(*string*) The user-specified name of the .dta file containing the combined input .dta files

**savecombdir**(*string*) The user-specified file path where the combined input .dta file (named in the savecombdta(string) option) is saved. If the directory is not specified along with “savecombdta(string)”, the combined input .dta file will be saved in the user’s working directory by default.

**savesumdta**(*string*) The user-specified name of the .dta file containing the summary CGM metrics.

**savesumdir**(*string*) The user-specified file path where the summary CGM metric dataset (named in the savesumdta(string) option) is saved. If the directory is not specified along with “savesumdta(string)”, the summary CGM metric dataset will be saved in the user’s working directory by default.

**saveplotdir**(*string*) The user-specified file path where any plots (histogram as specified in the “hist” option and CGM glucose tracings as specified in the “plot” option) are saved. If “saveplotdir(string)” is not specified, then plots are not saved.

## 3. Output

After running the **cgmstats** package which contains at minimum the variable names for ID, timestamp, and glucose measurement, the result is a Stata dataset containing the following CGM metrics (output variables):

| **Output variable** | **Description** |
| --- | --- |
| *first_glucose_reading* | The timestamp of the first CGM reading |
| *ndayswear* | Number of days CGM sensor was worn |
| *total_sensor_readings* | Total number of CGM sensor readings |
| *percent_cgm_wear* | The number of sensor readings as a percentage of possible obtained readings by the CGM (given time worn) |
| *mean_sensor\** | Mean of sensor glucose values |
| *median_sensor\** | Median sensor glucose value |
| *q1_sensor\** | First quartile sensor glucose value |
| *q3_sensor\** | Third quartile sensor glucose value |
| *min_sensor\** | Minimum of sensor glucose values |
| *max_sensor\** | Maximum of sensor glucose values |
| *sd_sensor\** | Standard deviation of sensor glucose values |
| *cv_sensor\** | Coefficient of variation (%) of sensor glucose values (100*SD/mean glucose) |
| *\*min_spent_X<sub>i</sub>_Y<sub>i</sub>* | Minutes spent in the target blood glucose range (X-Y, where X and Y are determined by any combination of cut points from options "hypo" and "hyper") |
| *\*percent_time_X<sub>i</sub>_Y<sub>i</sub>* | Minutes spent in the target blood sugar range (X-Y), as a percentage of the total time CGM was worn |
| *\*min_spent_under_X<sub>i</sub>* | Total number of minutes spent below cut point(s) X (i.e., 54 and 70 mg/dL; option "hypo") |
| *\*percent_time_under_X<sub>i</sub>* | Minutes spent below cut point(s) X (i.e., 54 and 70 mg/dL), as a percentage of the total time CGM was worn |
| *episodes_under_X<sub>i</sub>* | The count of glucose episodes below cut point(s) X (i.e., 54 and 70 mg/dL; option "hypo"), for user-defined time (i.e., default at least 30 minutes; option hypo_exc_lngth) |
| *avg_hypo_lngth_X<sub>i</sub>* | The mean length (in minutes) of episodes below cut point(s) X (i.e., 54 and 70 mg/dL; option "hypo"). Minimum length to be considered an episode defined by option "hypo_exc_lngth" (default 30 minutes). |
| *\*min_spent_over_X<sub>i</sub>* | Total number of minutes spent above cut point(s) X (i.e., 180 and 250 mg/dL; option "hyper") |
| *\*percent_time_over_X<sub>i</sub>* | Minutes spent above cut point(s) X (i.e., 180 and 250 mg/dL), as a percentage of the total time CGM was worn |
| *episodes_over_X<sub>i</sub>* | The count of glucose episodes above cut point(s) X (i.e., 180 and 250 mg/dL; option "hyper"), for user-defined time (i.e., default at least 30 minutes; option hyper_exc_lngth) |
| *avg_hyper_lngth_X<sub>i</sub>* | The mean length (in minutes) of episodes above cut point(s) X (i.e., 180 and 250 mg/dL; option "hyper"). Minimum length to be considered an episode defined by option "hyper_exc_lngth" (default 30 minutes). |
| *auc_over_X<sub>i</sub>* | Area under the sensor glucose curve over cut point(s) X (i.e., 180 and 250 mg/dL; option “hyper”), calculated using the trapezoidal rule |

\*These metrics are produced overall and for daytime and nighttime (as defined by options “daystart” and “dayend”)

If *X*_*i*_ contains a decimal point, the decimal point is converted to an underscore in the variable name. For example, if *X*_*i*_ is 2.9, the variable name would be “percent_time_under_2_9”.

The output dataset is saved in the specified working directory ($dir) and titled “cgm_summary_file”. When the option by(day) is used, the output dataset is titled “cgm_summary_file[day]”, i.e. “cgm_summary_file1” for day 1, “cgm_summary_file2” for day 2.

## 4. Example 1

This example (dataset “cgm_example1.dta”) inputs a CGM dataset from a system that recorded glucose every 5 minutes (vs. the default of 15 minutes), denoted using the option freq(5). Here, the user includes CGM readings occurring any time before 12pm on 2/6/2022 [timebefore(“2022/02/06 12:00”)]. The example also specifies that the CGM readings were measured in mmol/L [option unit(mmol/L)] and the summary statistics are outputted in mmol/L.

~~~
cgmstats, id(id) glucose(GlucoseValue) ///
time(GlucoseDisplayTime) ///
dtadir(‘c(pwd)’/example1) freq(5) unit(mmol/L) ///
timebefore(“2022/02/06 12:00”) ///
hist(mean_sensor cv_sensor percent_time_3_9_10 ///
percent_time_over_10, freq) plot(3.9 10)///
savecombdta(cgmcombined) ///
savesumdta(cgm_summary_fille) saveplotdir(‘c(pwd)’/example1)
~~~

Note that the decimal point “3.9” (one of the default values for “hypo” when units are mmol/L and “hypo” values are not specified) is converted to an underscore in the output variable name, “percent_time_3_9_10”. Stata does not accept decimal points in variable names.

This is a snapshot of the Stata output dataset:

~~~
list id first_glucose_reading ndayswear mean_sensor in 1/10
~~~

|  | id | first_glucose_reading | ndayswear | mean_sensor |
| --- | --- | --- | --- | --- |
| 1. | 1 | 2017/12/03 05:31 | 5.1 | 8.8 |
| 2. | 2 | 2018/06/06 10:49 | 6.3 | 6.6 |
| 3. | 3 | 2018/06/06 05:02 | 6.5 | 7.2 |
| 4. | 4 | 2018/02/24 11:31 | 6.3 | 11.5 |
| 5. | 5 | 2018/01/30 12:09 | 6.2 | 8.6 |
| 6. | 6 | 2018/03/10 09:36 | 5.8 | 8.5 |
| 7. | 7 | 2018/03/11 05:23 | 2.6 | 7.3 |
| 8. | 8 | 2018/03/13 06:43 | 6.5 | 7.0 |
| 9. | 9 | 2018/03/10 17:39 | 7.0 | 7.0 |
| 10. | 10 | 2018/02/28 11:41 | 6.3 | 10.3 |

**Figure 1.**
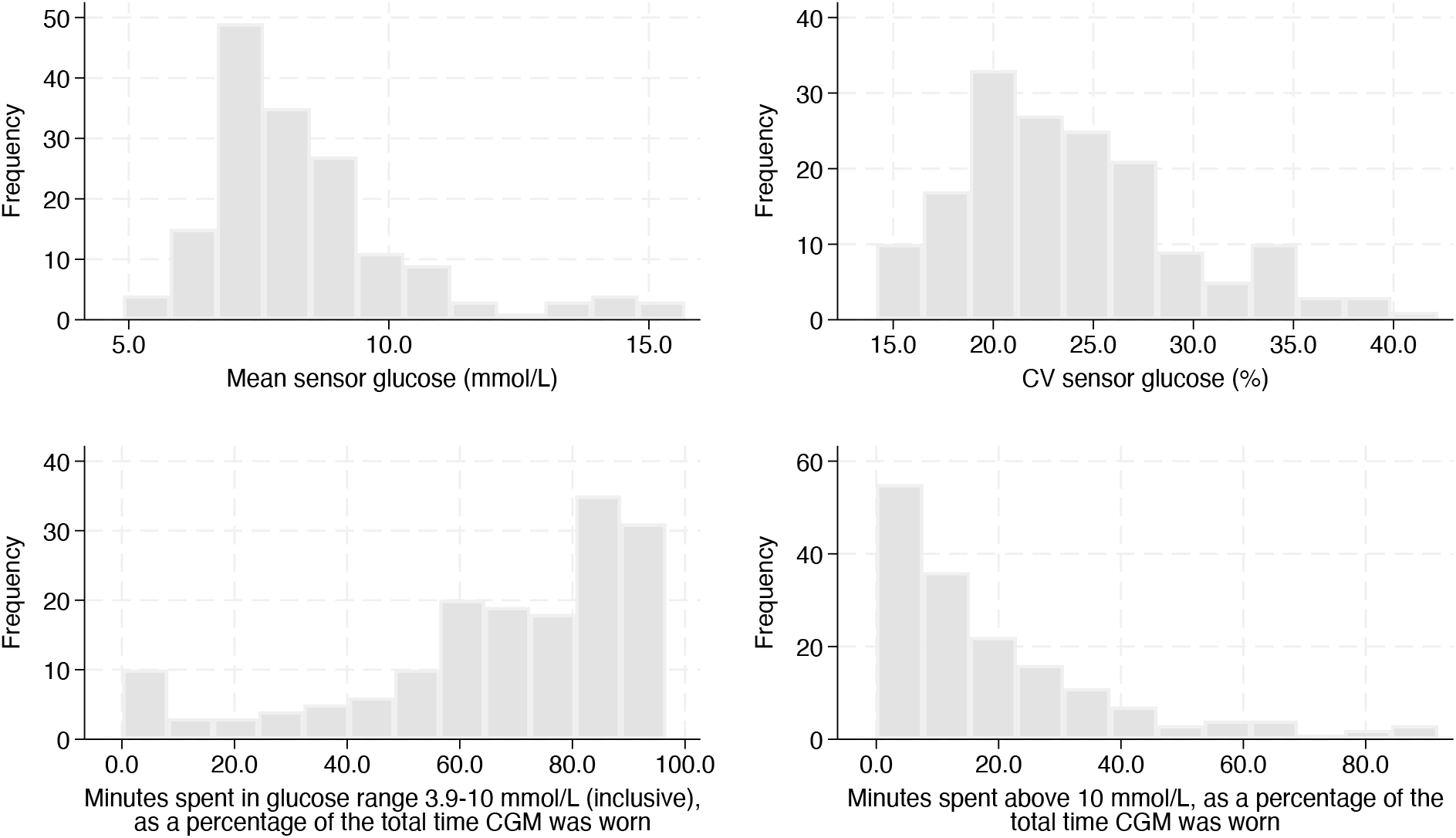
Frequency histograms of CGM specified summary statistics. Note: Statistics are specified in “hist” option.

The CGM glucose tracings for each individual in the dataset are output using the option “plot(3.9 10)”. Below are the tracings for the first 2 individuals in the dataset.

**Figure 2.**
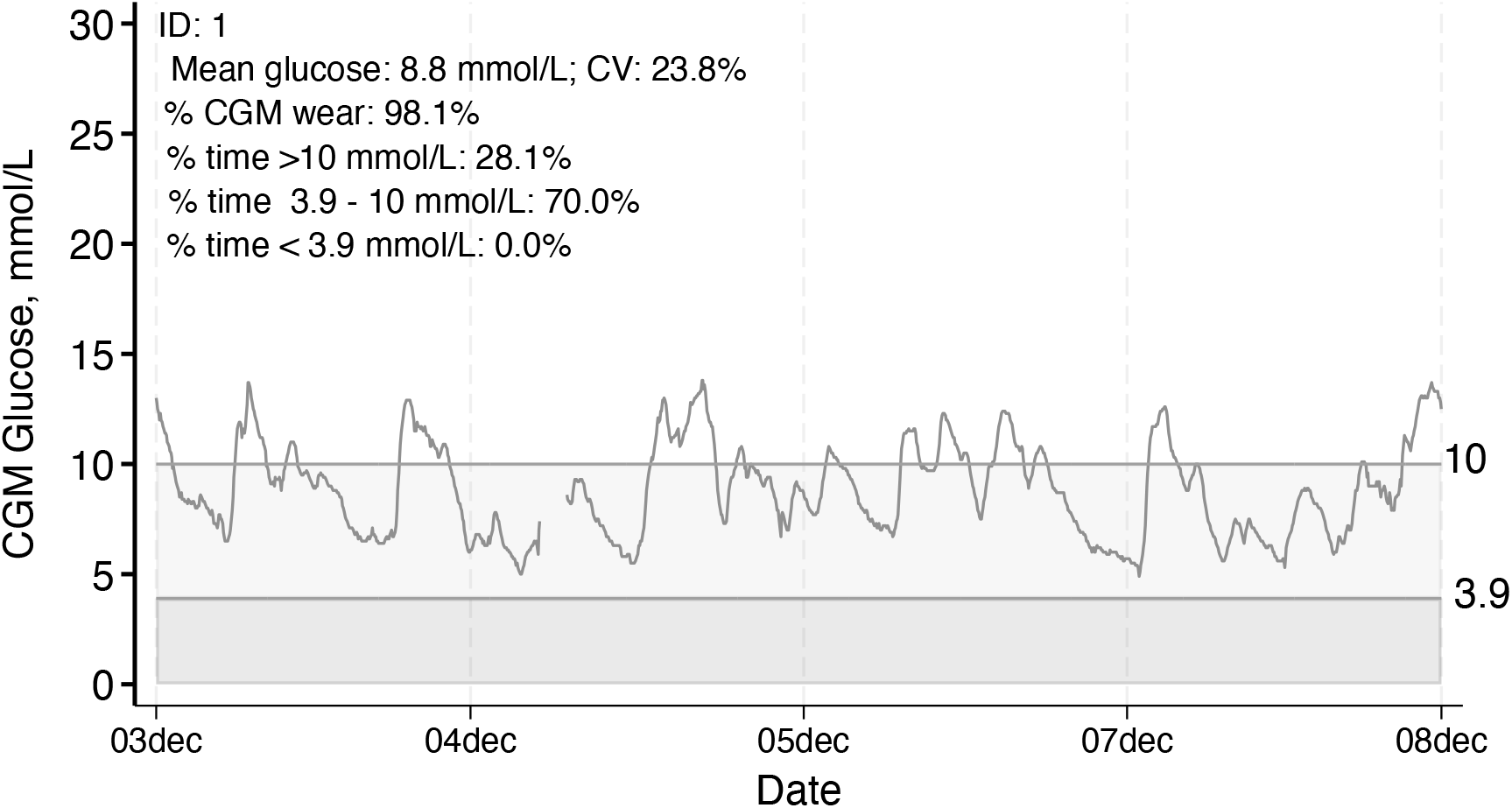
Individual CGM glucose tracing with summary data displayed, Example 1, ID 1.

**Figure 3.**
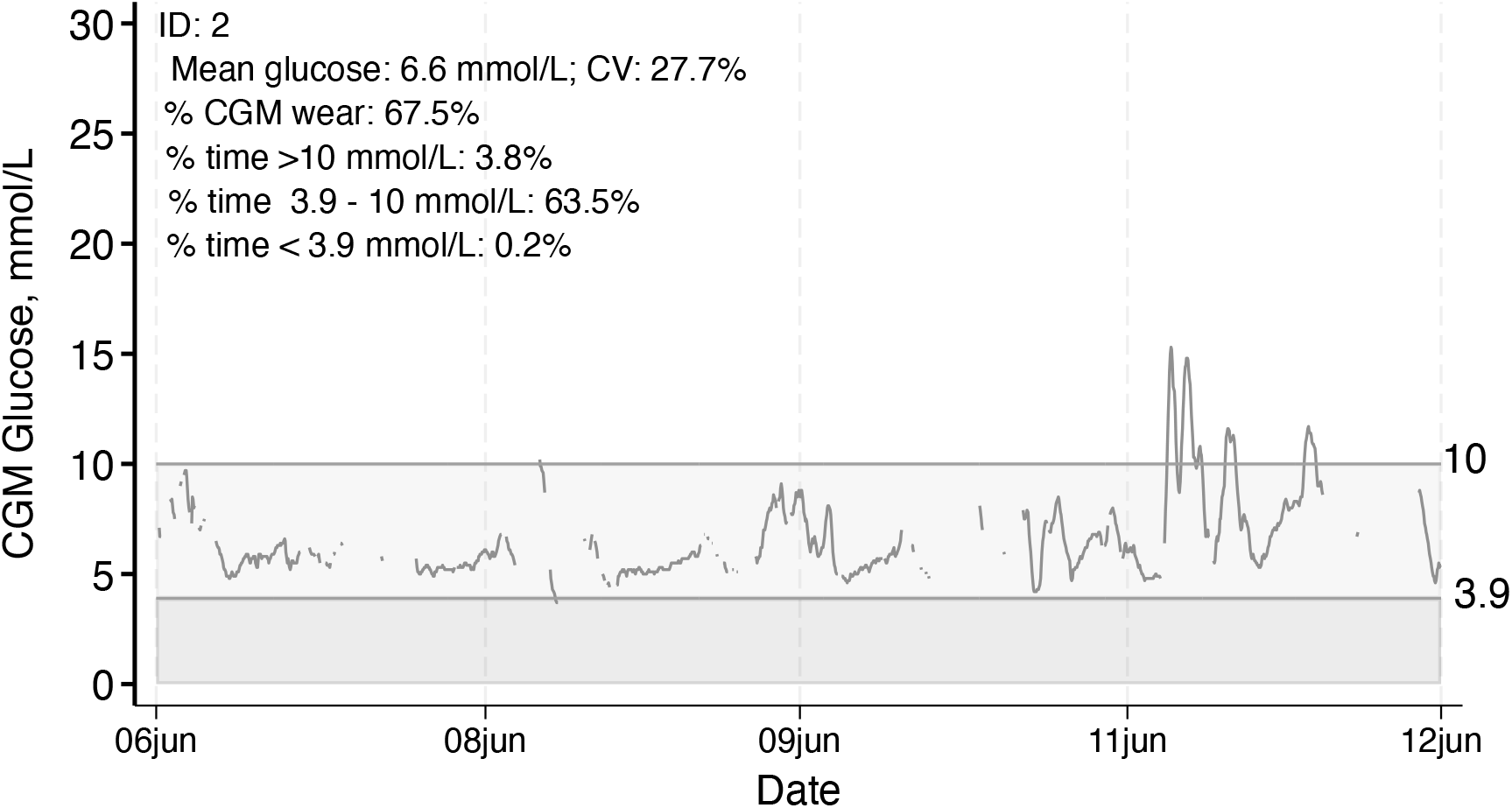
Individual CGM glucose tracing with summary data displayed, Example 1, ID 2.

The lighter shading indicates time in range and the darker shading indicates time below range, as specified by the user or according to the default (time in range 70-140 mg/dL, or 3.9-7.8 mmol/L).

In the example CGM tracings above, there are gaps between CGM glucose readings. If we specify the option “fill” [e.g. cgmstats, id(id) glucose(GlucoseValue)

time(GlucoseDisplayTime) dtadir(‘c(pwd)’/example1) freq(5) timebefore(“2022/02/06 12:00”) unit(mmol/L) plot(3.9 10, **fill**) hist(mean_sensor cv_sensor percent_time_3_9_10 percent_time_over_10, freq) savecombdta(cgmcombined)

savesumdta(cgm_summary_file) saveplotd9r(‘c(pwd)’/example1], these gaps are filled using linear interpolation.

**Figure 4.**
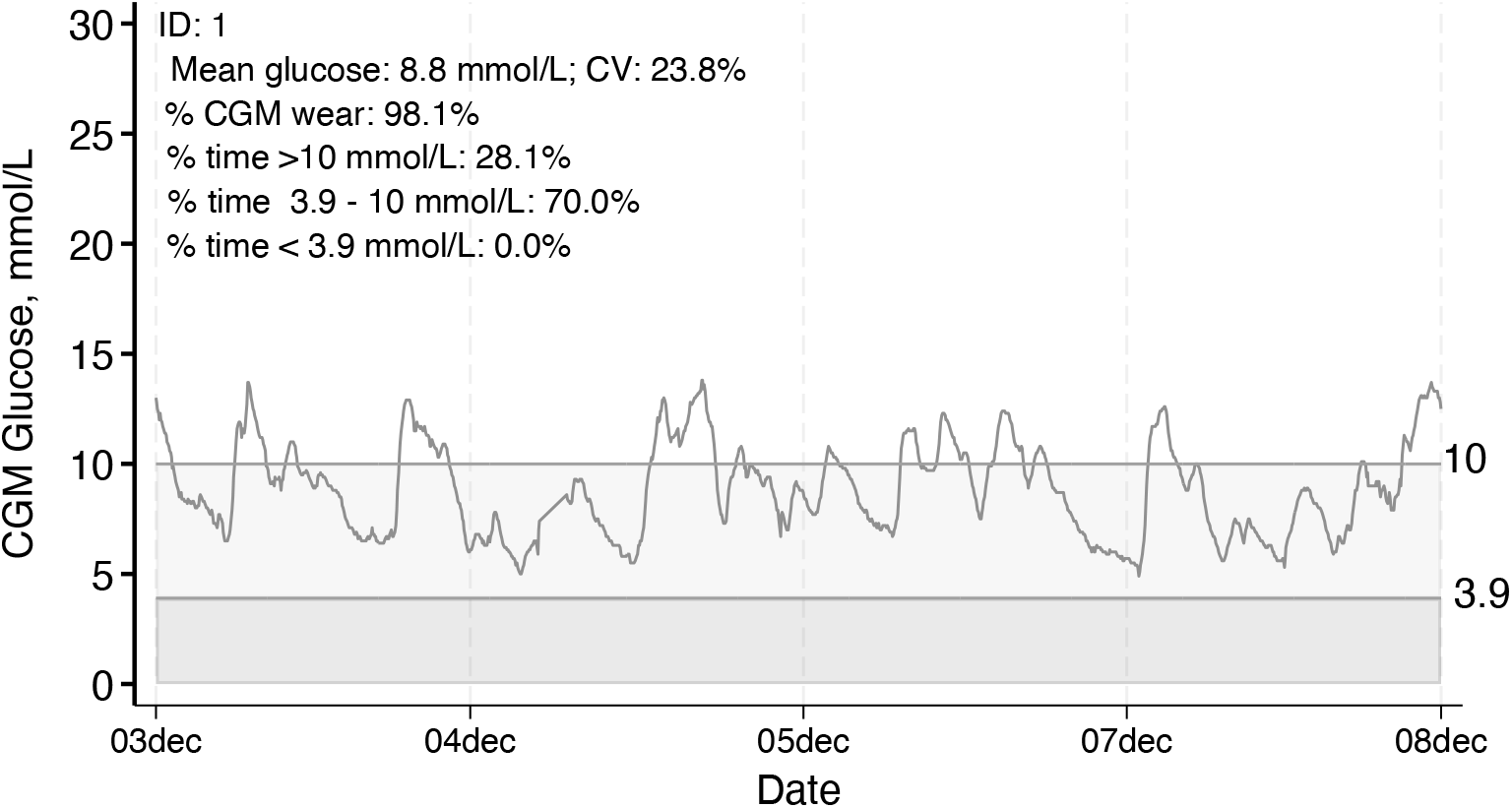
Individual CGM glucose tracing with summary data displayed and gaps filled using linear interpolation, Example 1, ID 1.

**Figure 5.**
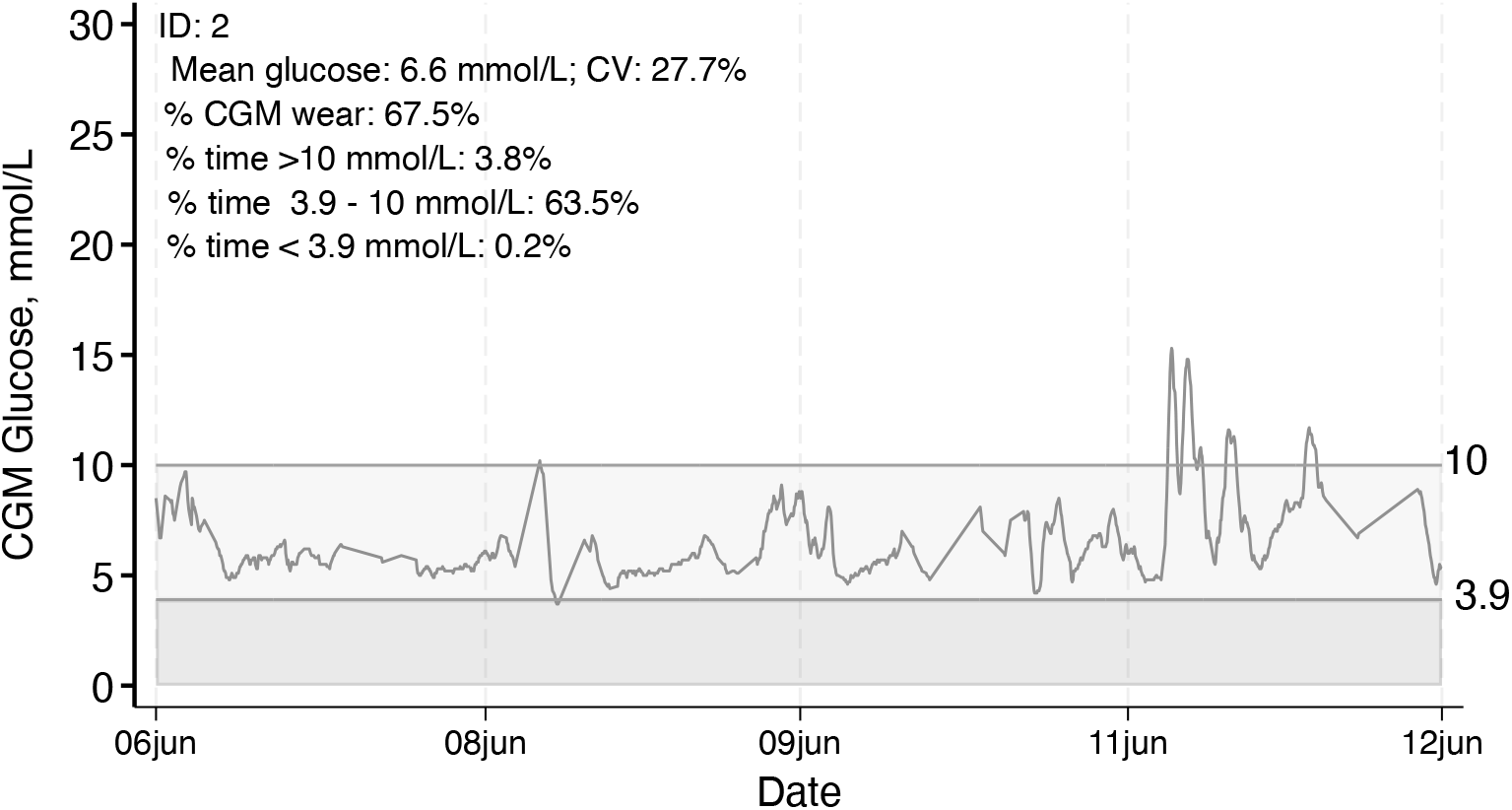
Individual CGM glucose tracing with summary data displayed and gaps filled using linear interpolation, Example 1, ID 2.

In both versions, the summary statistics printed on the plots are from the actual data (no interpolation). Summary statistics can be suppressed using the option, “plot(3.9 10, nostats)”.

## 5. Example 2

This example (dataset “cgm_example2.dta”) specifies that CGM summary metrics are output in one dataset by ID and by day (i.e., multiple rows per person) using the option [by(id day)]. In this example, we drop the first hour of CGM wear [firsthours(1)] and define hypoglycemic episodes as 15 minutes below 54 or 70 mg/dL and hyperglycemic episodes as 45 minutes above 140, 180, or 250 mg/dL.

~~~
cgmstats, id(subjectid) glucose(sensorglucose) ///
time(timestamp) dtadir(‘c(pwd)’/example2) by(id day) ///
hyper_exc_lngth(45) hypo_exc_lngth(15) firsthours(1) ///
plot(70 140)
~~~

This is a snapshot of the Stata output dataset:

~~~
list subjectid day total_sensor_readings mean_sensor in 1/20
~~~

|  | subjectid | day | total_sensor_readings | mean_sensor |
| --- | --- | --- | --- | --- |
| 1. | 1 | 1 | 32 | 95.6 |
| 2. | 1 | 2 | 5 | 85.6 |
| 3. | 1 | 3 | 94 | 90.5 |
| 4. | 1 | 4 | 96 | 94.0 |
| 5. | 1 | 5 | 96 | 86.9 |
| 6. | 1 | 6 | 96 | 91.9 |
| 7. | 1 | 7 | 96 | 86.7 |
| 8. | 1 | 8 | 96 | 88.5 |
| 9. | 1 | 9 | 96 | 92.8 |
| 10. | 1 | 10 | 96 | 96.8 |
| 11. | 1 | 11 | 96 | 92.0 |
| 12. | 1 | 12 | 96 | 89.6 |
| 13. | 1 | 13 | 53 | 84.1 |
| 14. | 1 | 14 | 72 | 97.1 |
| 15. | 1 | 15 | 55 | 83.1 |
| 16. | 2 | 1 | 38 | 97.3 |
| 17. | 2 | 2 | 96 | 100.8 |
| 18. | 2 | 3 | 96 | 99.7 |
| 19. | 2 | 4 | 96 | 99.9 |
| 20. | 2 | 5 | 96 | 96.7 |

Note: Histograms are not produced when option by(id day) is used.

The CGM glucose tracings for each person and each day of wear are output using the option “plot(70 140)”. Below are examples, showing the tracings for day 5 and 11 of an individual (subjectid=6) in the dataset.

**Figure 6.**
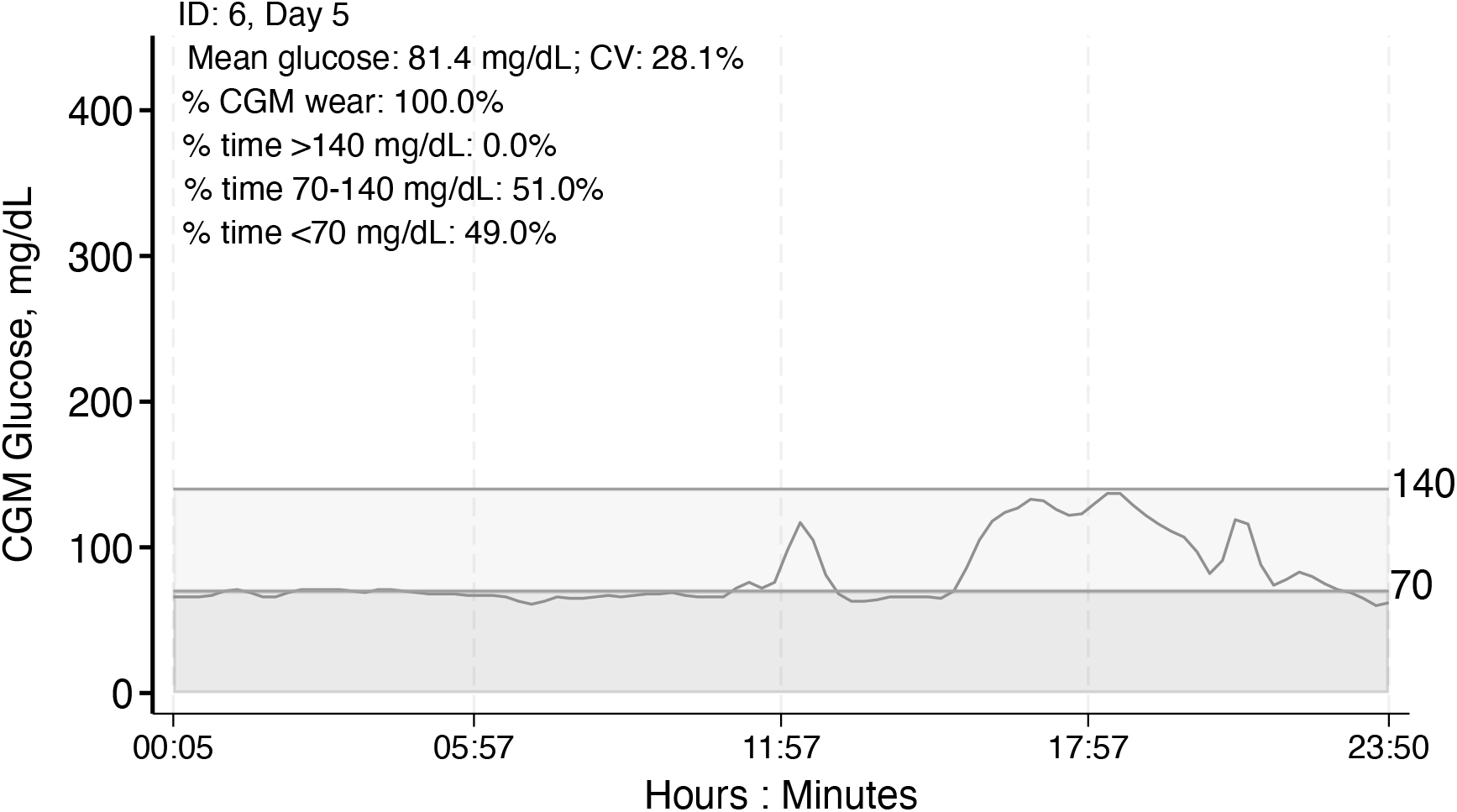
Individual CGM tracing by day with day specific summary statistics, Example 2, ID 6, Day 5.

**Figure 7.**
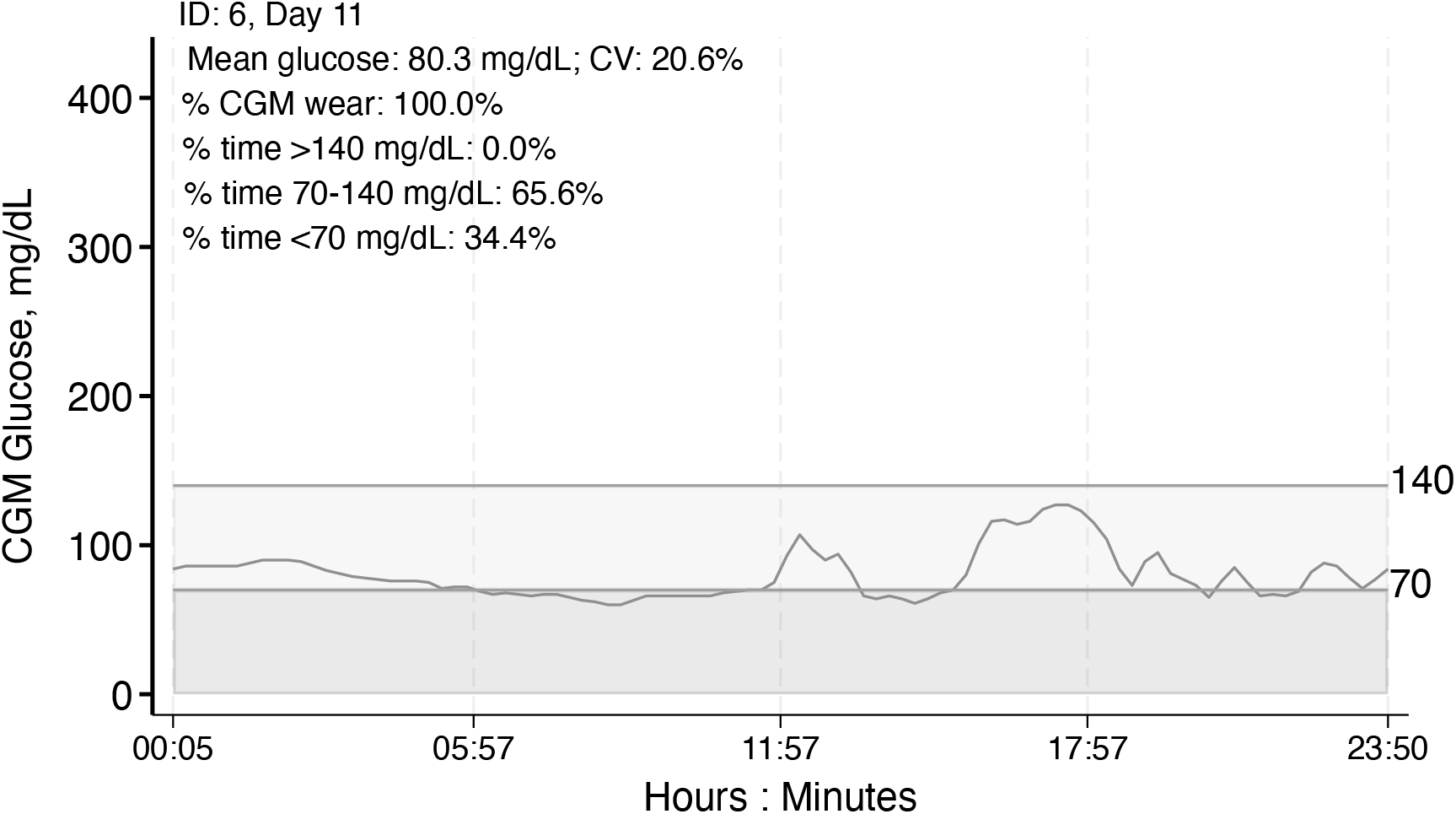
Individual CGM tracing by day with day specific summary statistics, Example 2, ID 6, Day 11.

## 6. Conclusions

This article introduced **cgmstats**, a Stata package that provides a flexible framework for processing CGM data and generating summaries and visualizations of data from several CGM systems. CGM systems are increasingly available and used in research studies and clinical trials. By streamlining workflows, **cgmstats** enhances rigor and reproducibility in CGM research and lowers barriers to incorporating CGM data into epidemiologic, clinical and translational research.

Emerging research using CGM data involves advanced approaches including machine learning (ML) models, for example (Dave et al. 2021; Klonoff et al. 2025). Although the **cgmstats** package focuses on descriptive statistics and visualizations, these are a critical first step for more complex analyses and this package can provide the input required for ML models and other advanced modeling techniques. Integrating CGM data with information from other wearable devices or applications, such as dietary logs and physical activity trackers, may also be of interest to researchers and can be facilitated by the output produced from this package.

The functions and flexibility offered in **cgmstats** provides Stata users with an efficient and automated method to process, summarize and visualize CGM data from multiple systems. This **cgmstats** package is well positioned to evolve alongside advances in digital health research, supporting broader applications and accelerating the next generation of high-quality scientific research using these clinically important data.

## Data Availability

All data produced in the present study are available upon reasonable request to the authors.

https://ideas.repec.org/c/boc/bocode/s459576.html

## 7 Acknowledgements

We thank the members of Dr. Elizabeth Selvin’s Diabetes Data group for their invaluable support with this work including pilot testing and feedback.

Dr. Selvin is supported by National Institutes of Health (NIH)/National Institute of Diabetes and Digestive and Kidney Diseases (NIDDK) grant R01 DK128837 and a Merit Award from the American Heart Association. Dr. Fang is supported by NIH/NIDDK career development award K01 DK138273.

## About the authors

Natalie Daya Malek is a Research Associate at the Johns Hopkins Bloomberg School of Public Health (Welch Center for Prevention, Epidemiology, and Clinical Research) and a longtime Stata user.

Dan Wang and Sui Zhang are Senior Biostatisticians at the Johns Hopkins Bloomberg School of Public Health (Welch Center for Prevention, Epidemiology, and Clinical Research) and longtime Stata users.

Michael Fang is an Assistant Professor at the Johns Hopkins Bloomberg School of Public Health (Welch Center for Prevention, Epidemiology, and Clinical Research).

Amelia Wallace is an Assistant Scientist at the Johns Hopkins Bloomberg School of Public Health (Welch Center for Prevention, Epidemiology, and Clinical Research) and longtime Stata user.

Scott Zeger is a professor at Johns Hopkins Bloomberg School of Public Health in the Department of Biostatistics.

Elizabeth Selvin is a professor at Johns Hopkins Bloomberg School of Public Health and the director of the Welch Center for Prevention, Epidemiology, and Clinical Research.

